# Prediction of One-Week Sport-Related Concussion Symptom Severity Using the Sport Concussion Assessment Tool and CogState Brief Battery

**DOI:** 10.64898/2026.02.12.26346072

**Authors:** Anna F. Butts, James W. Hickey, Gershon Spitz, Becca Xie, Lauren P. Giesler, Lauren J. Evans, Terence J. O’Brien, Sandy R. Shultz, Bradley J. Wright, Stuart J. McDonald, William T. O’Brien

**Affiliations:** Department of Behavioral Neuroscience, Oregon Health and Science University, Portland, OR, USA; Department of Neuroscience, School of Translational Medicine, Monash University, VIC, Australia; Department of Monash-Epworth Rehabilitation Research Centre, School of Psychological Sciences, Monash University, VIC, Australia; Department of Neurology, Alfred Health, The Alfred Hospital, VIC, Australia; Division of Medical Sciences, Faculty of Health, University of Victoria, BC, Canada; School of Psychology and Public Health, La Trobe University, VIC, Australia

**Keywords:** SRC, mTBI, biomarker, prognostic, diagnostic, SCAT, RPQ

## Abstract

**BACKGROUND:** The recovery from sport-related concussion (SRC) is highly heterogenous, with many individuals experiencing symptoms that persist beyond typical recovery timeframes. The early identification of individuals at risk of prolonged symptoms is therefore critical to inform timely interventions and set realistic recovery expectations. Although acute symptom burden is one predictor of future symptom burden, reliance on self-reported measures may limit objectivity and reduce clinical utility in settings where symptom evaluation may be unreliable.

In this prospective cohort study, we evaluated the discriminatory accuracy of the CogState Brief Battery, alone and in combination with the Sport Concussion Assessment Tool (SCAT), to classify Australian football players with SRC from Australian footballers without SRC at 24-hours post-injury/match. Furthermore, we examined whether CogState performance and symptom severity at 24 hours were associated with symptom outcomes at one-week post-injury.

Adult amateur Australian football players (n=181) were recruited following SRC (n=109 SRC, 86% male) or after a non-injured match (n=72, 90% male). Participants completed the CogState Brief Battery, SCAT and Rivermead Post Concussion Questionnaire (RPQ) at 24-hours and one-week post-injury or match. Area under the receiver operating characteristic (AUC) analyses quantified the ability of 24-hour CogState task performance and SCAT symptom severity to distinguish SRC from controls. Linear regression models examined associations between CogState performance and symptom severity (SCAT and RPQ), within and across the 24-hour and one-week time points. Additional models evaluated whether combining 24-hour symptom severity assessments with CogState performance improved prediction of one-week symptom burden and symptomatic status.

SCAT symptom severity demonstrated excellent discriminatory classification accuracy for SRC versus controls at 24-hours post-injury (AUC [95% CI]: 0.949 [0.916 - 0.981]). CogState task performance showed lower discriminatory accuracy but demonstrated fair classification and prognostic utility (e.g., Identification task AUC [95% CI]: 0.666 [0.582 - 0.750]). CogState performance at 24-hours was significantly associated with overall symptom severity at both 24-hours and one-week, as well as with symptom severity across individual symptom domains. In combined models, 24-hour symptom severity and CogState performance independently contributed to the prediction of symptomatic from asymptomatic individuals at one-week post-SRC (e.g., Identification task AUC [95% CI]: 0.721 [0.606 - 0.835] for classification based on <4 versus ≥4 symptoms).

These findings indicate that CogState performance at 24-hours post-SRC may serve as an objective adjunct to subjective symptom-based reporting, supporting both diagnosis and early prognostication in the clinical evaluation of SRC.

## 1. Introduction

Sport-related concussion (SRC) is a form of mild traumatic brain injury (mTBI) sustained in athletic settings. The Centers for Disease Control and Prevention estimates 1.6 to 3.8 million SRCs occur annually in the United States; however, this figure is likely underestimated given many athletes fail to recognize or report symptoms.^1–4^ SRC recovery is variable: while most athletes recover within two weeks, others experience persistent symptoms for weeks or months.^5–11^ Recognizing the risks associated with returning to sport before complete recovery, many sporting organizations now mandate the full resolution of symptoms and a minimum stand-down period, prior to return-to-play. Despite the development of diagnostic tools, there are limited measures that can accurately predict who will recover within the typical timeline and who will experience a protracted recovery.^12–17^ This gap limits clinician’s ability to guide early management decisions, set appropriate recovery expectations, and make informed return-to-play decisions.^4,9,10,12,14,18^

Acute symptom burden, defined as the severity and number of symptoms within the first 72 hours post-injury, has consistently emerged as a predictor of prolonged recovery.^7–10,12,19–22^ Self-reported symptom evaluation measures like the Sport Concussion Assessment Tool (SCAT) and the Rivermead Post-Concussion Symptoms Questionnaire (RPQ) are widely implemented and capture cognitive, emotional, somatic, and sleep-related symptoms.^12,15,23^ The SCAT is a structured symptom checklist developed to facilitate standardized evaluation of acute symptoms and includes a self-administered symptom rating and a brief cognitive assessment, which may only be conducted by a healthcare professional.^21,24,25^ While valuable tools, both the SCAT and RPQ rely on self-report measures, which may be influenced by external pressures to underreport, the non-specific nature of SRC symptoms, and limited sensitivity to subtle cognitive deficits within 72 hours of SRC.^14,23-29^

Combining self-report tools with computerized neurocognitive tests may improve the objectivity and classification accuracy of SRC assessment.^4,7,29-31^ One such tool, the CogState Brief Battery, objectively measures multiple cognitive domains such as attention, working memory, and psychomotor speed.^4,32,33^ Compared with other cognitive assessment tools used after SRC, the CogState battery has significant practical utilities: it does not require a healthcare professional to administer, can be carried out across multiple devices or online, and repeat tests are not prone to practice effects and can be overseen by different administrators, facilitating its potential utility in community settings.^14,30-34^ Integrating a strong, but subjective, tool such as self-reported symptom assessment scores, with an objective battery such as the CogState, may enhance the sensitivity for predicting protracted recovery and offer improved accuracy, brevity, and clinical feasibility for SRC diagnosis and management compared to each tool used in isolation.^7,13,18,32-36^ The potential added value of integrating subjective symptom measures with objective neurocognitive testing for SRC assessment and recovery prognostication warrants further investigation.

In this study, we assessed symptom burden using the SCAT and RPQ, and cognitive performance using the CogState battery at 24-hours and one-week post-injury in amateur Australian football players. The primary aim was to determine whether 24-hour CogState performance provides prognostic information regarding the presence and burden of symptoms at one-week post-injury, and whether these associations differed across symptom sub-domains. As a secondary aim, we examined which combinations of symptom and cognitive measures best discriminated athletes with SRC from controls in the acute period of injury. We hypothesized that self-reported symptom measures collected in a research setting would, when used in isolation, demonstrate the greatest classification accuracy for SRC and the strongest predictive utility for one-week symptom burden; however, we further hypothesized that this classification and predictive accuracy would improve when symptom measures were combined with CogState performance. Finally, we hypothesized that the predictive utility of the CogState battery would not be limited to cognitive symptom burden.

## 2. Methods

### 2.1 Participant Recruitment

Participants in this study were drawn from two cohorts (BIOREC 1^9^ and BIOREC 2) that underwent testing with the CogState Brief Battery at 24-hours, and one-week post-match/SRC.^9^ Recruitment was performed through the Victorian Amateur Football Association from April 10, 2021, to September 2, 2023. All protocols were reviewed and approved by the Monash University Human Research Ethics Committee.

Adult male and female Australian footballers with suspected SRC were referred to the research team by club medical staff (physiotherapists, sports trainers or doctors) present on matchday. Individuals with SRC were identified at the sideline by club medical staff using the Concussion Recognition Tool 5/6 or SCAT 5/6. Inclusion criteria for the SRC group were consistent with consensus definitions of SRC from the Concussion in Sport Group^12^ requiring i) a biomechanically plausible mechanism of injury, ii) at least one observable sign and/or two or more symptoms not better explained by confounding factors, iii) suspicion of SRC by club medical staff. Control participants were recruited through convenience sampling and featured a mixture of players that had participated in an Australian football match without sustaining an injury (i.e. healthy control) and players identified by club medical staff as having sustained a musculoskeletal injury in the preceding match. Inclusion criterion for the healthy control group was completion of an Australian football match without sustaining a concussion or musculoskeletal injury. Inclusion criterion for the musculoskeletal group was the presence of a musculoskeletal injury sustained during match play that required treatment by club medical staff initially predicted to result in 1-3 weeks of missed play. Exclusion criteria for all groups included sustaining a concussion in the preceding six months, a history of moderate or severe traumatic brain injury, a neurological or psychiatric condition that could significantly impact cognition, or current significant musculoskeletal injury.

### 2.2 Data collection

Testing was conducted at two time-points: 24-hours and one-week post-SRC/match. Written informed consent was obtained from all participants at the commencement of the first testing session. Demographic information (sex, age, years of education, body mass index, years of collision sport participation), concussion history, and SRC characteristics (loss of consciousness and/or memory impairment) were self-reported via written survey at the first time-point. Symptom evaluation and cognitive testing was conducted at the 24-hour and one-week testing sessions.

### 2.3 Measures

#### Sport Concussion Assessment Tool (SCAT) symptom checklist

The SCAT (Version 5) symptom checklist comprises 22 symptoms, each rated on a 7-point Likert Scale (0 = none, 6 = severe) based on the athlete’s self-report at the time of assessment. Total symptom severity scores range from 0 to 132 points.^25,37^ SCAT symptoms were classified into four domains as described by Simons et al. (2023)^23^: i) Somatic - ‘Headaches’, ‘Pressure in head’, ‘Neck pain’, ‘Nausea or Vomiting’, ‘Dizziness’, ‘Balance problems’, ‘Sensitivity to light’, ‘Sensitivity to noise’, and ‘Fatigue or low energy’; ii) Cognitive - ‘Feeling like “in a fog”, ‘Difficulty concentrating’, ‘Difficulty remembering’, and ‘Confusion’; iii) Emotional - ‘More emotional’, ‘Irritability’, ‘Sadness’, and ‘Nervous or anxious’; and iv) Sleep: -’Drowsiness’ and ‘Trouble falling asleep’.

#### Rivermead post-concussion symptoms questionnaire (RPQ)

The RPQ is a self-report evaluation comprising 16 mTBI-related symptoms rated on a 5-point Likert scale (0 = not experienced at all, 4 = a severe problem). This self-report asks participants to report the degree to which symptoms have been problematic in the preceding 24 hours compared to prior to their concussion. Scores of each of the 16 mTBI-related symptoms were added to give possible symptom severity scores from 0 to 52.^23,38^ RPQ symptoms were classified into four domains as described by Simons et al. (2023)^23^: i) Somatic - ‘Headaches’, ‘Feelings of dizziness’, ‘Nausea and/or vomiting’, ‘Noise sensitivity (easily upset by loud noise)’ and ‘Light sensitivity (easily upset by bright light)’; ii) Cognitive – ‘Forgetfulness, poor memory’; iii) Emotional - ‘Being irritable, easily angered’, ‘Feeling depressed or tearful’ and ‘Feeling frustrated or impatient’, and iv) Sleep - ‘Sleep disturbance).

#### CogState Brief Battery

The CogState battery consists of four computerized tasks (10–15 minute duration) measuring cognitive function of concussed athletes.^36^ The four cognitive tasks (Detection [DET], Identification [IDN], One-Back [ONB] and One-Card Learning [OCL]) involve playing cards, assessing the speed and accuracy traits of cognitive performance. Briefly, the Detection task is a reaction time test that assesses visual processing speed, in which athletes attend to the rule, “Has the card turned over?”; the Identification task is a choice reaction time test that assesses simple decision-making with the rule, “Is the card red?”; the One-Back task is a working memory test with the rule, “Was the previous card the same?”; and the One Card Learning task is a visual learning test with the rule, “Have you seen this card before in this task?” ^35,36,39,40^ Speed was the primary performance measure for the DET, IDN and ONB tasks. Accuracy was the primary measure for the OCL task.^40^

### 2.4 Statistical Analyses

Demographic data was compared using Chi-squared test for categorical data (sex and history of concussion) and Wilcoxon rank sum test for numerical data (age, body mass index, years of education, number of previous concussions and years of collision sport). As standard for the CogState, latency on the DET, IDN and ONB were log10 transformed and accuracy on the OCL was corrected using arcsine transformation of the square root of the proportion of correct responses. The classification accuracy of the SCAT symptom severity and the four CogState tasks at 24-hours and one-week post-SRC were assessed with logistic regressions (*ln*) and area under the receiver operating characteristic curve (AUC) analyses (*pROC*). Additional analyses evaluated the combined classification accuracy of the SCAT with each CogState task. As the RPQ requires symptoms to be assessed relative to pre-concussion status, RPQ data was only analyzed within the SRC group. Within SRC participants, linear regression models (*lm*) were used to examine associations between performance on the four CogState tasks at 24-hours and symptom severity scores (SCAT and RPQ) at the same timepoint. Standardized β-coefficients were estimated using the *parameters* package. Association between CogState tasks performance at 24-hours and symptom severity scores at one-week, as well as CogState tasks performance at one-week with symptom severity scores at the same timepoint were assessed using the same approach. To explore whether these relationships were driven by specific symptom domains, the models were repeated with the number of somatic, cognitive, emotional or sleep symptoms as the outcome variable. To assess whether early symptom severity alone or in combination with CogState performance predicted symptom severity at one-week, linear regression models were fit with one-week SCAT symptom severity as the outcome and 24-hour SCAT severity alone, or combined with each CogState task, as predictors. Analyses were performed similarly with RPQ symptom severity. To evaluate the ability of 24-hour SCAT symptoms and CogState performance to predict ongoing symptoms at one-week, logistic regression and AUC analyses were conducted. The discriminatory ability of 24-hour SCAT symptom severity, performance on each CogState task individually, and the combination of SCAT with each CogState task to distinguish symptomatic from asymptomatic individuals was assessed. To evaluate prediction of ongoing symptom burden, two symptom-count thresholds at one-week were used: ≥1 vs 0 (any symptom endorsement) and ≥4 vs <4, a more stringent definition selected a priori to reduce misclassification due to common non-specific symptoms frequently reported by uninjured athletes. All logistic and linear regressions were adjusted for age, sex and years of education. Analyses were two-tailed (p<0.05) and performed using RStudio Version 4.2.2 (R Foundation for Statistical Computing).

## 3. Results

### 3.1 Recruitment and participant demographics

This study included a total of 181 Australian football players with a median age of 23.0 (IQR: 5.2) years. The cohort was primarily male (89.0%). There were 109 individuals with SRC and 72 control individuals. Demographic variables were highly similar between the SRC and control groups for sex, age, years of education, and body mass index. There was a slightly higher self-reported history of concussion in the SRC group (66%) compared to the control group (50%, p=0.031), and a greater number of previous concussions in the SRC group (median [IQR]: 1.0 [3.0]) than the control group (median [IQR]: 0.5 [2.0], p=0.017). Although statistically different, the 24-hour testing session was performed at similar times between the SRC group (median [IQR]: 22.5 [7.3] hours, range: 16.0-51.5 hours) and control group (median [IQR]: 24.3 [5.8] hours, p=0.029, range: 17.3-73.0 hours). There was no difference between time of testing at one-week between the SRC group (median [IQR]: 8.6 [2.2] days, range: 6.62-12.10 days) and control group (median [IQR]: 8.2 [2.8] days, range: 7.67-11.0 days). Full demographic, concussion history and SRC characteristics are available in **Table 1**.

**Table 1.** Participant demographics and injury characteristics.

| **Characteristic** | **SRC<sup>a</sup>,**<br>**N = 109** | **Control<sup>a</sup>,**<br>**N = 72** | **p-value<sup>b</sup>** |
| --- | --- | --- | --- |
| **Sex** |  |  | 0.420 |
| *Female* | 15 (14.0%) | 7 (9.7%) |  |
| *Male* | 94 (86.0%) | 65 (90.0%) |  |
| **Age (years)** | 22.5 (5.2) | 23.6 (5.4) | 0.140 |
| **Years of Education** | 16.0 (2.0) | 16.0 (2.0) | 0.100 |
| **Body Mass Index** | 23.7 (2.4) | 24.4 (2.4) | 0.064 |
| **History of Concussion** |  |  | 0.031 |
| *Yes* | 72 (66.0%) | 36 (50.0%) |  |
| *No* | 37 (34.0%) | 36 (50.0%) |  |
| **Number of Previous Concussions** | 1.0 (3.0) | 0.5 (2.0) | 0.02 |
| **Years of Collision Sport** | 13.0 (5.0) | 14.0 (6.0) | 0.53 |
| **24h Session Time post-injury/match (hours)** | 25.5 (7.3) | 24.3 (5.8) | 0.029 |
| **1w Session Time post-injury/match (days)** | 8.6 (2.2) | 8.2 (2.8) | >0.99 |
| **Loss of Consciousness** |  |  |  |
| *Yes* | 32 (29.0%) |  |  |
| *No* | 70 (64.0%) |  |  |
| *Unsure* | 7 (6.4%) |  |  |
| **Memory Impairment** |  |  |  |
| *Yes* | 48 (44.0%) |  |  |
| *No* | 61 (56.0%) |  |  |

*A small subset had missing data, i.e. Years of education (control: n=1), years of collision sport participation (SRC: n=1; control: n=1), 24h session time post-injury/match (SRC: n=6; control: n=1), 1w session time post-injury/match (SRC: n=4; control: n=7). All data except for 24h Session and 1w Session Time post-injury/match is self-reported.*
<sup>a</sup>n (%); Median (IQR)
<sup>b</sup>Pearson's Chi-squared test; Wilcoxon rank sum test; Fisher's exact test

### 3.2 The classification accuracy of the SCAT and CogState tasks for SRC

Firstly, we investigated the classification accuracy of the SCAT and CogState tasks for SRC in Australian footballers at 24-hours and one-week post-injury. The 24-hour SCAT severity demonstrated excellent classification accuracy for SRC versus control (AUC = 0.949, 95% CI [0.916 - 0.981]) (**Table 2**). Although lower than that of the SCAT, 24-hour performance on the four CogState tasks demonstrated fair classification accuracy for SRC versus control, with the highest observed for the DET task (AUC = 0.683, 95% CI [0.596 - 0.769]). When combined with 24-hour SCAT, CogState tasks provided minimal improvement in classification accuracy.

**Table 2.**
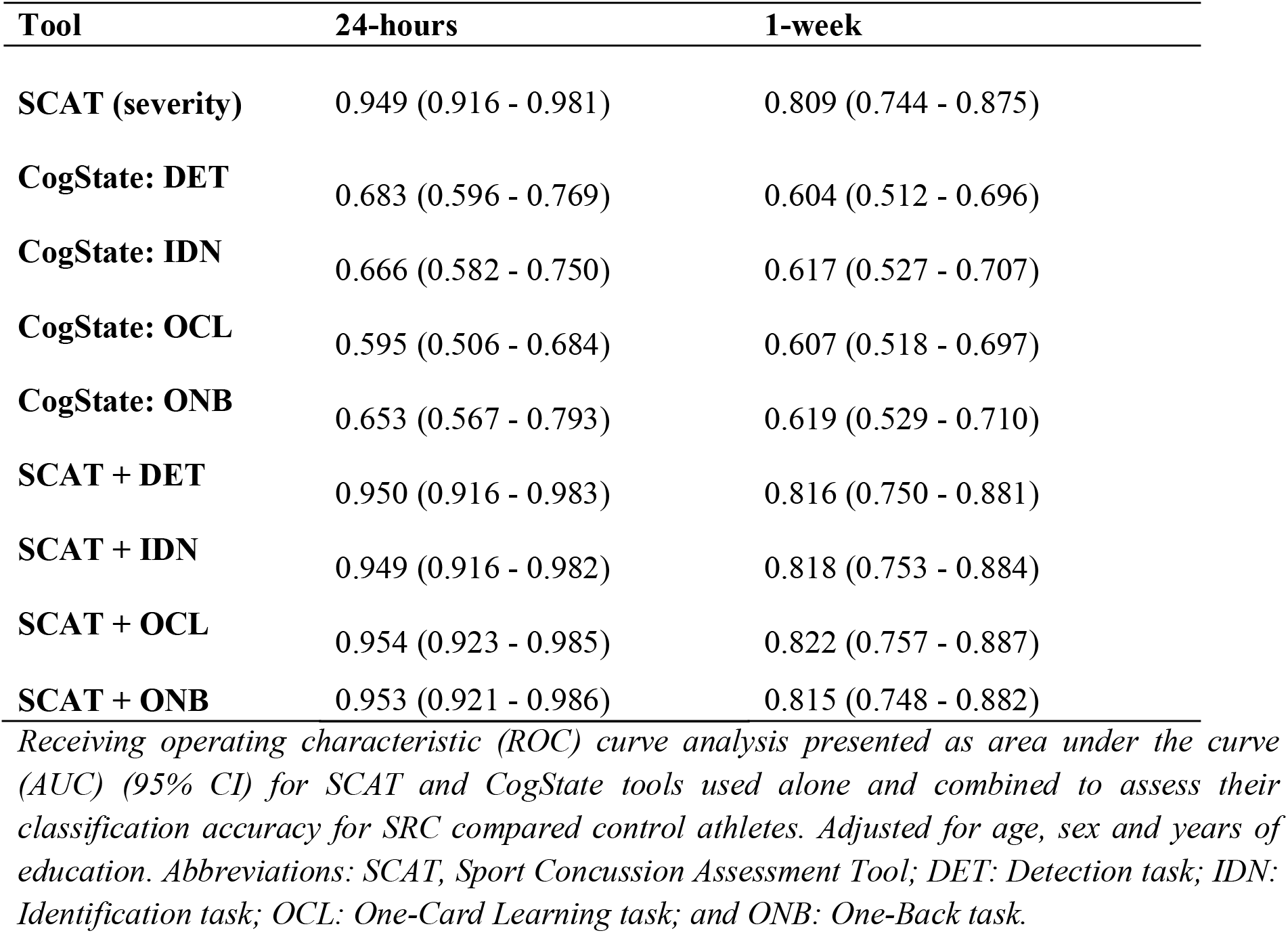
AUC analyses of symptom severity and CogState tests for discriminating athletes with SRC at 24-hours and one-week post-injury (SRC) / post-match (controls).

At one-week post-injury, SCAT symptom severity showed reduced accuracy compared to 24-hours but maintained a good classification accuracy for SRC versus control (AUC = 0.809, 95% CI [0.744 - 0.875]) (**Table 2**). Each of the four CogState tasks demonstrated similar performance (DET: AUC = 0.604, 95% CI [0.512 - 0.696]; IDN: AUC = 0.617, 95% CI [0.527 - 0.707]; OCL: AUC = 0.607, 95% CI [0.518 - 0.697]; ONB: AUC = 0.619, 95% CI [0.529 - 0.710]) and provided little to no added benefit when combined with the SCAT. Full statistical results are available in **Table 2**.

### 3.3 CogState tasks as predictors for 24-hour and one-week symptom severity

Linear regression models (adjusted for age, sex and years of education) were used to examine the relationship between CogState performance at 24-hours with self-reported symptoms at 24-hours and one-week (Table 3). At 24-hours, slower reaction times on the DET, IDN and ONB tasks were associated with greater symptom severity on both SCAT and RPQ. Additionally, poorer accuracy on the OCL task was associated with a greater symptom severity score on the RPQ, but not the SCAT. The largest effect size was observed for 24-hour DET performance with 24-hour SCAT (β-coefficient: 0.46, 95% CI [0.25, 0.68]), followed by RPQ symptom severity (β-coefficient: 0.45, 95% CI [0.24, 0.66]).

**Table 3.** Linear regression model for CogState tasks as predictors of SRC symptoms.

|  | **24-hour SCAT** | **24-hour RPQ** | **1-week SCAT** | **1-week RPQ** |
| --- | --- | --- | --- | --- |
| **24-hour DET** | 0.46 [0.25, 0.68]<br>p < 0.001 | 0.45 [0.24, 0.66]<br>p < 0.001 | 0.36 [0.14, 0.59]<br>p = 0.002 | 0.20 [-0.03, 0.44]<br>p = 0.093 |
| **24-hour IDN** | 0.44 [0.24, 0.65]<br>p < 0.001 | 0.39 [0.18, 0.60] p<br>< 0.001 | 0.38 [0.17, 0.60]<br>p < 0.001 | 0.28 [0.05, 0.50]<br>p = 0.016 |
| **24-hour OCL** | 0.05 [-0.17, 0.27]<br>p = 0.670 | -0.27 [-0.48, -0.07]<br>p = 0.011 | -0.12 [-0.34, 0.10]<br>p = 0.277 | 0.14 [-0.09, 0.36]<br>p = 0.223 |
| **24-hours ONB** | 0.29 [0.08, 0.50]<br>p = 0.010 | 0.27 [0.07, 0.48]<br>p = 0.010 | 0.24 [0.03, 0.45]<br>p = 0.023 | 0.25 [0.04, 0.46]<br>p = 0.023 |
| **1-week DET** |  |  | 0.29 [0.10, 0.49]<br>p = 0.004 | 0.30 [0.10, 0.50]<br>p = 0.004 |
| **1-week IDN** |  |  | 0.42 [0.23, 0.60]<br>p < 0.001 | 0.35 [0.15, 0.54]<br>p < 0.001 |
| **1-week OCL** |  |  | -0.21 [-0.40, -0.01]<br>p = 0.036 | -0.05[-0.25, 0.15]<br>p = 0.620 |
| **1-week ONB** |  |  | 0.31 [0.12, 0.50]<br>p = 0.002 | 0.27 [0.07, 0.47]<br>p = 0.008 |

*Linear regression of overall symptoms and CogState data is presented as standardized β-coefficient (95% CI) and p-value at 24-hours post-injury and one-week post-injury. Adjusted for age, sex and years of education. Abbreviations: SCAT, Sport Concussion Assessment Tool; RPQ, Rivermead Post-Concussion Symptoms Questionnaire; DET, CogState Detection task; IDN, CogState Identification task; OCL, CogState One-Card Learning task; and ONB, CogState One-Back task.*

Subsequently, we evaluated the ability of 24-hour CogState tasks to predict symptom burden (SCAT and RPQ) at one-week post-injury. Slower reaction times on DET, IDN, and ONB tasks were similarly associated with greater symptom severity on the SCAT, and the IDN and ONB were associated with greater symptom severity on the RPQ, although these relationships were characterized by smaller effect sizes than the corresponding within 24-hour analyses. OCL performance at 24-hours showed no significant association with symptom severity on either measure at one-week.

Furthermore, similar associations were observed between one-week CogState performance and one-week symptom severity, with slower reaction times on the DET, IDN and ONB tasks, again associated with a greater symptom severity on both the SCAT and RPQ. Poorer accuracy on the OCL task was also associated with greater SCAT, but not RPQ symptom severity. Notably, the effect sizes of these associations were relatively lower than those observed between 24-hour CogState performance and 24-hour or one-week symptom burden. Full statistical results are provided in **Table 3**.

### 3.4 CogState tasks as predictors of symptom sub-domain severity at 24-hours at one-week

To determine whether CogState performance was predictive of the presence and/or severity of cognitive symptoms specifically, or extended to other symptom sub-domains, linear regression models were used to assess the relationship between CogState performance with the severity of i) somatic, ii) emotional, iii) cognitive and iv) sleep-related symptoms at 24-hours and one-week post-injury (Supplementary Tables 1-4).

Within both the 24-hour and one-week timepoints, performance on the CogState DET, IDN, and ONB tasks were consistently associated with all symptom domains on both the SCAT and RPQ at the same timepoint. Furthermore, 24-hour IDN task performance was predictive of one-week SCAT severity across the sub-domains. In line with the overall analysis, the DET, IDN and ONB tasks were predictive of one-week cognitive and emotional symptom severity. Conversely, SCAT somatic symptoms were only predicted by DET and IDN performance, and sleep-related SCAT symptoms were only predicted by IDN performance. No CogState task was predictive of RPQ symptom severity for these domains. Full statistical details are available in Supplementary Tables 1-4.

### 3.5 Predictive utility of 24-hour symptom severity and CogState performance for one-week symptom severity

Symptom severity at 24-hours on both the SCAT (*β*-coefficient: 0.61, 95% CI [0.47, 0.75]) and RPQ (*β*-coefficient: 0.47, 95% CI [0.32, 0.62]) were significant predictors of one-week symptoms (quantified with the SCAT and RPQ respectively). When performance on the each of the CogState tasks were added to the linear regression, although the *β*-coefficients were lower than that of the 24-hour symptom severity, the IDN, OCL, and ONB tasks were additionally found to be significant predictors of one-week SCAT symptom severity. Similarly, performance on the OCL and ONB tasks were found to be significant predictors of one-week RPQ symptom severity in a model including 24-hour RPQ symptoms. Full results are available in **Table 4**.

**Table 4.** Linear regression models comparing symptom severity alone and in combination with CogState to predict one-week post-injury SCAT and RPQ symptom severity (separate models)

| Model | Variable | Coefficient [95% CI] | p-value | R<sup>2</sup> |
| --- | --- | --- | --- | --- |
| **24h SCAT** | 24h SCAT | **0.61 [0.47, 0.75]** | **< 0.001** | **0.374** |
| **24h SCAT + DET** | 24h SCAT | **0.55 [0.40, 0.70]** | **< 0.001** | **0.391** |
|  | 24h DET | 0.14 [0.00, 0.29] | 0.057 |  |
| **24h SCAT + IDN** | 24h SCAT | **0.53 [0.38, 0.68]** | **< 0.001** | **0.398** |
|  | 24h IDN | **0.17 [0.02, 0.33]** | **0.024** |  |
| **24h SCAT + OCL** | 24h SCAT | **0.63 [0.49, 0.77]** | **< 0.001** | **0.393** |
|  | 24h OCL | **-0.14 [-0.28, 0.00]** | **0.049** |  |
| **24h SCAT + ONB** | 24h SCAT | **0.57 [0.43, 0.72]** | **< 0.001** | **0.387** |
|  | 24h ONB | **0.12 [-0.02, 0.27]** | **0.010** |  |
| **24 RPQ** | 24h RPQ | **0.47 [0.32, 0.62]** | **< 0.001** | 0.222 |
| **24h RPQ + DET** | 24h RPQ | **0.44 [0.27, 0.60]** | **< 0.001** | 0.229 |
|  | 24h DET | 0.09 [-0.07, 0.26] | 0.271 |  |
| **24h RPQ + IDN** | 24h RPQ | **0.41 [0.24, 0.57]** | **< 0.001** | 0.243 |
|  | 24h IDN | 0.16 [-0.01, 0.32] | 0.060 |  |
| **24h RPQ + OCL** | 24h RPQ | **0.51 [0.36, 0.66]** | **< 0.001** | **0.283** |
|  | 24h OCL | **0.25 [0.10, 0.40]** | **0.001** |  |
| **24h RPQ + ONB** | 24h RPQ | **0.41 [0.25, 0.57]** | **< 0.001** | 0.259 |
|  | 24h ONB | **0.20 [0.04, 0.36]** | **0.013** |  |

*Linear regression models for 24-hours post-injury symptoms assessed via SCAT and RPQ, and CogState performance as predictors of one-week post-injury symptoms. Adjusted for age, sex and years of education. Abbreviations: 24h, 24-hours post-injury, SCAT, Sport Concussion Assessment Tool; RPQ, Rivermead Post-Concussion Symptoms Questionnaire; DET, CogState Detection task; IDN, CogState Identification task; OCL, CogState One-Card Learning task; and ONB, CogState One-Back task.*

### 3.6 The predictive ability of SCAT and CogState for symptomatic vs asymptomatic individuals at one-week

Having determined that combining CogState and SCAT symptom assessments at 24-hours was predictive of symptom burden at one-week, we assessed the classification accuracy of 24-hour symptom severity and CogState performance in isolation and combined, for symptomatic versus asymptomatic individuals at one-week using i) ≥1 vs 0 and ii) ≥4 vs <4 thresholds (Table 5).

**Table 5.**
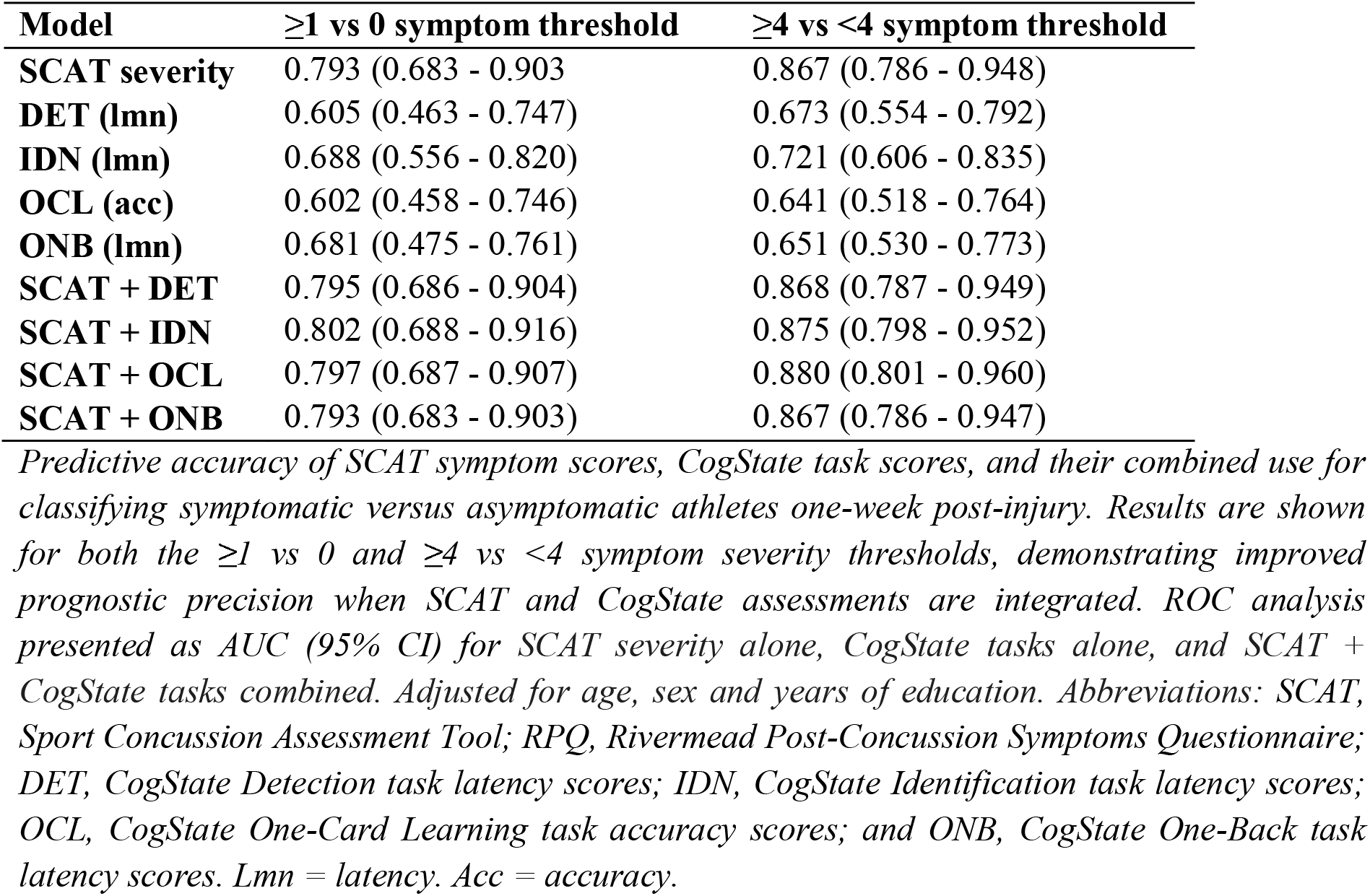
The predictive utility of 24-hour SCAT severity and CogState performance to predict symptomatic from asymptomatic individuals at one-week.

Using the ≥1 vs 0 symptom severity score threshold, we found that the symptom severity score from SCAT results demonstrated moderate predictive accuracy for symptomatic versus asymptomatic individuals with SRC (AUC = 0.793, 95% CI [0.683 - 0.903]). The CogState tasks had a fair classification accuracy, with the greatest accuracy observed for the IDN task (AUC = 0.688, 95% CI [0.556 - 0.82]). The addition of each CogState task to SCAT symptoms had minimal added benefit compared to SCAT symptoms alone. Results were highly similar for the ≥4 vs <4 symptom threshold; however, classification accuracies were substantially higher for all predictors with the SCAT demonstrating excellent utility (AUC = 0.867, 95% CI [0.786 - 0.948]) and the IDN task demonstrating a moderate accuracy (AUC = 0.721, 95% CI [0.606 - 0.835]).

## 4. Discussion

This study evaluated the utility of acute symptom burden, measured by the SCAT and RPQ, and objective cognitive performance, measured by the CogState battery, for predicting short-term symptom outcomes following SRC in amateur Australian football players. Secondary analyses examined how well these measures discriminated SRC cases from non-concussed control athletes. The key finding of this study was that for athletes with SRC, objective CogState testing conducted at 24-hours post-SRC was predictive of symptom burden at one-week. As expected, self-reported symptom burden at 24-hours consistently demonstrated the strongest ability to discriminate SRC from controls, whereas 24-hour CogState performance showed only moderate classification accuracy. Together, these findings indicate that the CogState battery offers complementary, objective information that may support the assessment, diagnosis, and prognosis of SRC, particularly in cases where self-reported symptoms cannot be relied upon in isolation.

### 4.1 The classification accuracy of the CogState battery for sport-related concussion

The management of SRC is inherently challenging because it relies heavily on self-reported symptoms. Although SRC is defined by its acute symptomology, exclusive reliance on symptom reports is problematic: the symptoms are often non-specific, individuals may lack insight into their presence or severity until after recovery, and some may hide or minimize symptoms to expedite return to work, sport or duty.^2,8,9,11^ Given that SRC is a clinical diagnosis defined by the presence of signs and symptoms, and considering that our cohort reported symptoms independently to their clinical management, it is unsurprising that self-reported symptoms were an excellent classifier of SRC. This likely reflects the upper limit of performance that symptom assessments can achieve in classifying SRC.

Although of lower magnitude than symptom assessment, the fair classification accuracy observed for the CogState Brief Battery, particularly the IDN task, suggests that this tool may provide some utility in distinguishing SRC from non-SRC cases. Previous studies have reported similar findings, with reaction time-based tasks demonstrating modest but meaningful sensitivity for classifying concussion from controls in the days following injury.^6,10,11,15,20,27^ Notably, our finding that the addition of CogState task performance to symptom assessment did not substantially improve the classification accuracy of the SCAT indicates that this battery is unlikely to detect additional cases beyond those identified through accurate symptom assessment alone. Furthermore, while the lower overall accuracy means that the CogState battery should not replace symptom evaluations, the objective behavioral information provided by this test could be used alongside symptom assessments to strengthen clinical decision-making, particularly in settings where symptoms may be ambiguous or under-reported.

### 4.2 Early CogState performance as a predictor of symptom resolution

The management of SRC is primarily guided by the trajectory of symptom resolution. For example, in Australian community sports, return-to-play guidelines mandate a minimum stand-down period of 21 days;^18,41^ however, athletes must be asymptomatic by seven days to return within this minimum timeframe.^9^ Furthermore, although early intervention is known to improve outcomes after concussion, clinical trials have been limited by recruitment challenges. With the majority of individuals spontaneously recovering within two weeks, trials require large sample sizes to distinguish normal recovery from improvements attributable to intervention.^4,9,11,40^ As a result, many studies recruit only those with ‘prolonged recovery’, necessitating a waiting period after injury before treatment begins, potentially missing a critical therapeutic window. This underscores the need for objective tools capable of identifying individuals at risk of prolonged symptoms acutely after injury.

Our finding that CogState performance, particularly the IDN task, had moderate classification accuracy in differentiating symptomatic (defined as four or more symptoms) compared to asymptomatic (fewer than four symptoms) individuals suggests that early cognitive testing may help identify those not likely to be symptom-free at one-week post-injury. Notably, although classification accuracy declined when a criterion of zero symptoms was applied (which may be confounded by symptoms unrelated to SRC), all tasks maintained moderate to good predictive utility. Given that community sport athletes who remain symptomatic at seven days are required to fulfil a longer-than-minimum stand-down period, an early predictive tool for symptom presence at this time-point could have considerable utility to manage player and team expectations regarding recovery and return-to-play timeframes. While further work is needed to validate these findings using context-dependent symptomatic thresholds, these results suggest that the CogState battery may serve as an objective tool to aid in the prediction of future symptom burden, enabling more targeted monitoring and potentially informing early, individualized interventions following SRC.

### 4.3 The association between CogState performance with symptom severity and symptom domains

We found that DET, IDN, and ONB tasks were all significant predictors of one-week SCAT symptom severity, both in isolation and in combination with 24-hour SCAT symptoms. Although CogState performance did not provide additional classification benefit beyond symptoms alone for identifying SRC and symptomatic individuals, the severity of symptoms at one-week was best explained by a model that includes both 24-hour SCAT symptom severity and IDN task performance. Further investigation into these relationships is warranted, particularly to determine whether there is a linear association between CogState performance and symptom severity, or whether pathological cut-offs can be identified. Nonetheless, these findings provide preliminary evidence that CogState testing could be used as an objective test alongside symptom evaluation to improve the prognostication of the severity of symptoms at one-week post-injury.

To further examine whether the relationship between 24-hour CogState performance and one-week symptoms was driven by specific symptom domains, we categorized and quantified symptom severity across cognitive, somatic, emotional, and sleep disturbance sub-domains. We found highly similar results to the overall analyses for somatic and emotional symptoms, suggesting that the CogState performance is sensitive to these domains in addition to the severity of self-reported cognitive symptom severity. In contrast, we saw comparatively fewer associations between 24-hour CogState performance and one-week sleep-related symptoms. While this may indicate a weaker relationship between CogState symptoms and sleep disturbances, it may also reflect limitations in the specificity of self-reported sleep symptoms, particularly in the acute setting. More targeted assessments, such as sleep diaries, wearable devices, or polysomnography may offer more reliable insight into sleep disturbances following SRC.^43,44^ Together, these findings suggest that acute CogState performance may provide an additional objective and prognostic tool to help inform somatic and emotional symptom trajectories, supporting individualized management following SRC.

### 4.4 Limitations

Several factors should be considered when interpreting the results of this study. Firstly, assessments were conducted only at 24 hours, and one week post-match/SRC timepoints, precluding evaluation of longer-term SCAT and CogState recovery trajectories, including time to symptom resolution. However, it is well-established that acute symptom burden predicts persistent symptoms.^7,8,10,15,20,45,46^ Secondly, despite known sex-related differences in concussion presentation and recovery,^10,39,40^ this study featured a comparatively small sample size of female athletes, preventing analyses into sex-specific differences. Finally, although symptom data were collected using standardized and routinely utilized forms in a research setting where athletes were unlikely to under-report symptoms, self-report measures remain inherently subjective. Future studies incorporating larger and more diverse cohorts, extended follow-up periods, and objective measures of recovery are warranted.

## 5. Conclusion

Here, we report that the CogState Brief Battery performance, which assessed psychomotor, attention, visual learning, and working memory, at 24-hours post-SRC demonstrates fair utility in prognosticating symptomatic from asymptomatic individuals at one-week post-injury in Australian football players who have sustained an SRC, as well as diagnosing players with SRC versus those without. Although self-reported symptoms collected in a research setting provided the strongest classification metrics, performance on the CogState battery, particularly the IDN task, may serve as objective behavioral data to support symptom assessment and strengthen clinical decision-making following SRC. Importantly, we found that 24-hour CogState performance added predictive value beyond symptom severity alone in predicting one-week symptom severity, suggesting its potential utility in assisting the prognostication of persistent symptoms, informing early interventions and setting realistic expectations for symptom trajectories post-SRC.

## Data Availability

All data produced in the present study is available upon reasonable request to the corresponding author.

## Acknowledgements

The authors thank the Victorian Amateur Football Association staff, clubs and players for their support of this study.

## Authors’ Contributions

A.F.B : Conceptualization; investigation; writing - original draft preparation; writing - review and editing. J.W.H. : Investigation; writing - review and editing. G.S. : Investigation; writing - review and editing. B.X. : Investigation; writing - review and editing. L.P.G. : Investigation; writing - review and editing. T.J.O. : Funding acquisition; resources; writing - review and editing. S.R.S. : Conceptualization; resources; supervision; writing - review and editing. B.J.W. : Conceptualization; writing - review and editing. S.J.M. : Conceptualization, investigation; resources; supervision; writing - review and editing. W.T.O. : Conceptualization; formal analyses; investigation; supervision; writing - original draft preparation; writing - review and editing. All authors have reviewed and approved this manuscript.

## Ethical Considerations

This study was approved by the Monash University Human Research Ethics Committee approval no. 27685 (approved March 15, 2021), and 37021 (approved March 21, 2023).

## Consent to Participate

All participants provided written informed consent prior to participating.

