## Supplemental Tables for "Prediction of One-Week Sport-Related Concussion Symptom Severity Using the Sport Concussion Assessment Tool and CogState Brief Battery"

**Supplementary Tables:**

**Supplementary Table 1: Linear regression model for CogState tasks as predictors of somatic SRC symptoms**

|  | **24 hours SCAT** | **24 hours RPQ** | **1-week SCAT** | **1-week RPQ** |
| --- | --- | --- | --- | --- |
| **24 hours DET** | 0.31 [0.08, 0.54]  p = 0.008 | 0.38 [0.16, 0.60]  p < 0.001 | 0.30 [0.07, 0.54]  p = 0.011 | 0.09 [-0.15, 0.33]  p = 0.450 |
| **24 hours IDN** | 0.33 [0.11, 0.54]  p = 0.003 | 0.34 [0.13, 0.55]  p = 0.002 | 0.31 [0.09, 0.53]  p = 0.007 | 0.10 [-0.13, 0.33]  p = 0.377 |
| **24 hours OCL** | -0.03 [-0.25, 0.19]  p = 0.801 | -0.39 [-0.59, -0.19]  p < 0.001 | -0.16 [-0.38, 0.06]  p = 0.162 | 0.15 [-0.07, 0.37]  p = 0.173 |
| **24 hours ONB** | 0.23 [0.02, 0.44]  p = 0.032 | 0.20 [-0.01, 0.41]  p = 0.057 | 0.18 [-0.04, 0.39]  p = 0.104 | 0.09 [-0.12, 0.31]  p = 0.396 |
| **1-week DET** |  |  | 0.25 [0.05, 0.45]  p = 0.014 | 0.15 [-0.06, 0.35]  p = 0.160 |
| **1-week IDN** |  |  | 0.37 [0.18, 0.56]  p < 0.001 | 0.18 [-0.02, 0.38]  p = 0.081 |
| **1-week OCL** |  |  | -0.21 [-0.41, -0.02]  p = 0.034 | -0.05 [-0.25, 0.15]  p = 0.601 |
| **1-week ONB** |  |  | 0.24 [0.05, 0.44]  p = 0.016 | 0.19 [-0.01, 0.38]  p = 0.066 |

Linear regression somatic symptom and CogState data is presented as standardized β-coefficient (95% CI) and p-value at 24 hours post-injury and one week post-injury. Adjusted for age, sex and years of education. Abbreviations: SCAT, Sport Concussion Assessment Tool; RPQ, Rivermead Post-Concussion Symptoms Questionnaire; DET, CogState Detection task; IDN, CogState Identification task; OCL, CogState One-Card Learning task; and ONB, CogState One-Back task.

**Supplementary Table 2: Linear regression model for CogState tasks as predictors of cognitive SRC symptoms**

|  | **24 hours SCAT** | **24 hours RPQ** | **1-week SCAT** | **1-week RPQ** |
| --- | --- | --- | --- | --- |
| **24 hours DET** | 0.46 [0.25, 0.68]  p < 0.001 | 0.42 [0.20, 0.64]  p < 0.001 | 0.33 [0.11, 0.56]  p = 0.005 | 0.20 [-0.03, 0.44]  p = 0.091 |
| **24 hours IDN** | 0.47 [0.27, 0.67]  p < 0.001 | 0.41 [0.20, 0.62]  p < 0.001 | 0.35 [0.13, 0.57]  p = 0.002 | 0.32 [0.10, 0.54]  p = 0.005 |
| **24 hours OCL** | 0.15 [-0.07, 0.37]  p = 0.184 | -0.28 [-0.49, -0.07]  p = 0.011 | -0.05 [-0.27, 0.17]  p = 0.65 | 0.10 [-0.12, 0.33]  p = 0.359 |
| **24 hours ONB** | 0.30 [0.10, 0.51]  p = 0.005 | 0.23 [0.02, 0.44]  p = 0.032 | 0.19 [-0.03, 0.40]  p = 0.084 | 0.24 [0.02, 0.45]  p = 0.030 |
| **1-week DET** |  |  | 0.21 [0.01, 0.41]  p = 0.038 | 0.30 [0.10, 0.50]  p = 0.004 |
| **1-week IDN** |  |  | 0.35 [0.16, 0.54]  p < 0.001 | 0.41 [0.22, 0.60]  p < 0.001 |
| **1-week OCL** |  |  | -0.15 [-0.35, 0.04]  p = 0.122 | -0.02 [-0.23, 0.18]  p = 0.814 |
| **1-week ONB** |  |  | 0.26 [0.06, 0.45]  p = 0.010 | 0.29 [0.10, 0.49]  p = 0.003 |

Linear regression cognitive symptom and CogState data is presented as standardized β-coefficient (95% CI) and p-value at 24 hours post-injury and one week post-injury. Adjusted for age, sex and years of education. Abbreviations: SCAT, Sport Concussion Assessment Tool; RPQ, Rivermead Post-Concussion Symptoms Questionnaire; DET, CogState Detection task; IDN, CogState Identification task; OCL, CogState One-Card Learning task; and ONB, CogState One-Back task.

**Supplementary Table 3: Linear regression model for CogState tasks as predictors of emotional SRC symptoms**

|  | **24 hours SCAT** | **24 hours RPQ** | **1-week SCAT** | **1-week RPQ** |
| --- | --- | --- | --- | --- |
| **24 hours DET** | 0.46 [0.25, 0.68]  p < 0.001 | 0.50 [0.30, 0.71]  p < 0.001 | 0.39 [0.16, 0.61]  p = 0.001 | 0.30 [0.06, 0.53]  p = 0.013 |
| **24 hours IDN** | 0.36 [0.15, 0.57]  p = 0.001 | 0.33 [0.12, 0.54]  p = 0.002 | 0.37 [0.15, 0.59]  p = 0.001 | 0.36 [0.14, 0.58]  p = 0.001 |
| **24 hours OCL** | 0.12 [-0.10, 0.33]  p = 0.286 | -0.09 [-0.31, 0.12]  p = 0.395 | -0.14 [-0.36, 0.08]  p = 0.216 | 0.10 [-0.12, 0.32]  p = 0.381 |
| **24 hours ONB** | 0.19 [-0.03, 0.40]  p =0.085 | 0.20 [-0.01, 0.41]  p = 0.060 | 0.32 [0.11, 0.53]  p = 0.003 | 0.37 [0.17, 0.58]  p < 0.001 |
| **1-week DET** |  |  | 0.33 [0.13, 0.53]  p = 0.001 | 0.35 [0.16, 0.54]  p < 0.001 |
| **1-week IDN** |  |  | 0.38 [0.19, 0.58]  p < 0.001 | 0.36 [0.16, 0.55]  p < 0.001 |
| **1-week OCL** |  |  | -0.14 [-0.34, 0.06]  p = 0.155 | -0.06 [-0.26, 0.14]  p = 0.559 |
| **1-week ONB** |  |  | 0.32 [0.13, 0.51]  p = 0.001 | 0.27 [0.07, 0.46]  p = 0.007 |

Linear regression emotional symptom and CogState data is presented as standardized β-coefficient (95% CI) and p-value at 24 hours post-injury and one week post-injury. Adjusted for age, sex and years of education. Abbreviations: SCAT, Sport Concussion Assessment Tool; RPQ, Rivermead Post-Concussion Symptoms Questionnaire; DET, CogState Detection task; IDN, CogState Identification task; OCL, CogState One-Card Learning task; and ONB, CogState One-Back task.

**Supplementary Table 4: Linear regression model for CogState tasks as a predictor of sleep symptoms**

|  | **24 hours SCAT** | **24 hours RPQ** | **1-week SCAT** | **1-week RPQ** |
| --- | --- | --- | --- | --- |
| **24 hours DET** | 0.39 [0.17, 0.61]  p < 0.001 | 0.39 [0.18, 0.61]  p < 0.001 | 0.19 [-0.05, 0.43]  p = 0.113 | 0.12 [-0.12, 0.36]  p = 0.320 |
| **24 hours IDN** | 0.42 [0.21, 0.62]  p < 0.001 | 0.38 [0.17, 0.58]  p < 0.001 | 0.31 [0.08, 0.53]  p = 0.008 | 0.21 [-0.01, 0.43]  p = 0.062 |
| **24 hours OCL** | -0.08 [-0.29, 0.14]  p = 0.493 | -0.19 [-0.40, 0.02]  p = 0.081 | -0.08 [-0.31, 0.15]  p = 0.485 | 0.09 [-0.12, 0.31]  p = 0.395 |
| **24 hours ONB** | 0.29 [0.09, 0.50]  p = 0.006 | 0.21 [0.00, 0.41]  p = 0.049 | 0.18 [-0.04, 0.39]  p = 0.109 | 0.21 [0.00, 0.42]  p = 0.056 |
| **1-week DET** |  |  | 0.24 [0.04, 0.45]  p = 0.019 | 0.31 [0.11, 0.50]  p = 0.003 |
| **1-week IDN** |  |  | 0.32 [0.12, 0.52]  p = 0.002 | 0.30 [0.11, 0.50]  p = 0.002 |
| **1-week OCL** |  |  | -0.16 [-0.36, 0.04]  p = 0.117 | -0.01 [-0.21, 0.19]  p = 0.915 |
| **1-week ONB** |  |  | 0.22 [0.02, 0.42]  p = 0.031 | 0.26 [0.06, 0.45]  p = 0.010 |

Linear regression sleep symptom and CogState data is presented as standardized β-coefficient (95% CI) and p-value at 24 hours post-injury and one week post-injury. Adjusted for age, sex and years of education. Abbreviations: SCAT, Sport Concussion Assessment Tool; RPQ, Rivermead Post-Concussion Symptoms Questionnaire; DET, CogState Detection task; IDN, CogState Identification task; OCL, CogState One-Card Learning task; and ONB, CogState One-Back task.
